# Beware of UVC sanitizers: not all are good

**DOI:** 10.1101/2020.09.19.20192757

**Authors:** Justo Arines, Carmen Bao-Varela

## Abstract

Disinfection is now one of the major concerns of society. Soap, or hydroalcohol are two known ways of sanitizing different objects. Now we can find in the market more sophisticated systems, which provide more confidence on their sanitizing effects, or that allows disinfection of objects that cannot be sanitized with soap or hydroalcoholic solutions. In this work we analyze a commercially available sanitizing device that claims the use of UVC radiation. We will show that this device is not suitable for complete disinfection of the objects introduced on the cabin. We provide spectral characterization of the sources, as well as irradiance measurements inside the cabin. We conclude that the cabin cannot be used for sanitizing most of the objects that are included in the documentation of the device.

## 1. Introduction

In these days, disinfection of daily used objects, like keys or mobile phones has increased significantly due to the concern on being infected with SARS-CoV-2. Added to soap or hydroalcoholic solutions, more sophisticated devices have been used, as ozone machines or UVC sterilizing cabins. A fast search in the internet provides a plethora of commercial disinfection UVC cabins of different sizes, materials, and prizes. From this variety, we highlight the product oriented to the disinfection of personal equipment, mobile phones, keys, rings… currently available in local shops, or in electronic commerce. These devices promise to disinfect the objects placed inside in a determined interval of time by using UVC radiation. Some of them, even pledge to disinfect in all directions, killing bacteria, fungi and virus.

UVC or UV germicidal radiation comprises de wavelength range (100 nm-280 nm) [1,2]. At these wavelengths, the photons present energy enough to be absorbed by the DNA or RNA bases limiting the capability of replication of microorganisms. Out of this range, it is also possible to inactivate microorganisms. For example, in the case of bacteria, radiation named violet-blue (380–480 nm) induce the creation of reactive oxygen species due to the absorption of photons by porphyrin molecules [3].

Disinfection not only depends on the wavelength but also on the dose (irradiance x time), which also depends on the pathogen [4–6]. Recent works performed using UVC on SARS-CoV-2 showed various doses. The work of Hiroko Inagaki [7] proposes a dose of 37 mJ/cm^2^ using UVC of 280 nm. The work of Andrea Bianco [8] proposes a dose of 6.7 mJ/cm^2^ using a source of 254 nm. Other work performed by Christiane Heilingloh [9] employing highly concentrated virus solution suggest a dose close to 1 J/cm^2^ to completely inactive the virus.

Studies on inactivation of microorganisms using UVB and UVA show even higher doses. In reference [10] Schuit et. al. concluded that with a dose around 3800 mJ/cm^2^ of UVA it is possible to achieve a reduction of the 90% of SARS-CoV-2. Concerning other microorganisms Rezaie et.al. [11] analyses the response to UVA. As an example, they show that for inactivating Escherichia Coli a dose of 3600 mJ/cm^2^ is needed. On the other hand, in the work of Tomb et al. [12] concerning the wavelength range known as violet-blue (380 – 500 nm) we can find a review of different doses depending on the microorganism. In this review we could not find microorganism inactivation with doses inferior to 4 J/cm^2^.

Recently, due to Covid-19 pandemic, and the need of masks disinfection, different studies have been published showing UVC cabin disinfection systems [13–14] and the relevance of the cabin design for providing uniform irradiance distribution on the object to be disinfected.

In the present manuscript we analyze a commercial disinfection cabin. In section 2 we present the experimental setup. Section 3 is devoted to results, while in section 4 we discuss the results. Finally, in section 5 we present the conclusions.

## 2. Experimental setup

The sanitizing chamber is a “UV sterilizer” with reference number 19025 distributed in Spain by H91 Investment S.L. bought in a local mall (see fig. 1). It is sold for disinfecting, mobile phones up to 7 inches, keys, rings… It claims a 360º object sterilization. The manual indicates a working wavelength of 253 nm. The time for disinfection is 5 minutes. The manual advices that the shadowed areas of the objects placed inside the chamber might not be disinfected. To operate the cabin, first we put the object inside, secondly close the cover, and press the Sterilize button. Important to notice that the cabin can be operated with the cover open, no security interlock is present in the system.

**Fig.1.**
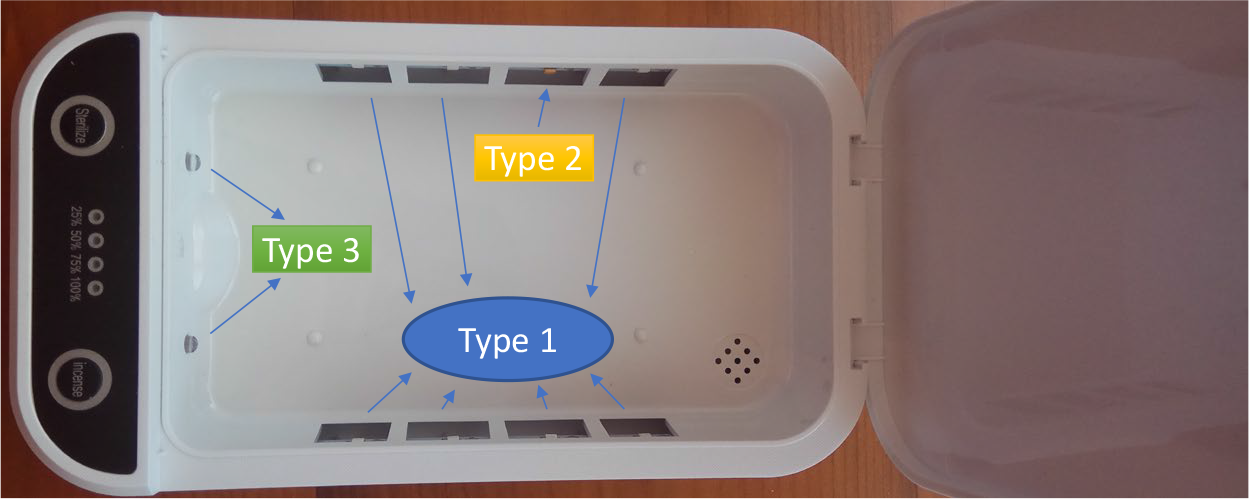
Sanitizing cabin with 3 types of LEDs

In figure 1 we show a picture of the sanitizing cabin. The inner part used for disinfection is 17.8 cm x 10.8 cm. It presents 10 LEDs, 7 of type 1, 1 of type 2, and 2 of type 3 (in next section we show their emission spectra). Only type 2 LED emits in the UVC region.

Figure 2 shows the cabin working with a model of a mobile phone and the spectrum probe. We use a melamine perforated plate to simulate a mobile model. The holes in the model are for placing the spectrometer probe in different parts of the mobile model, in order to measure the irradiance in its inferior part.

**Fig.2.**
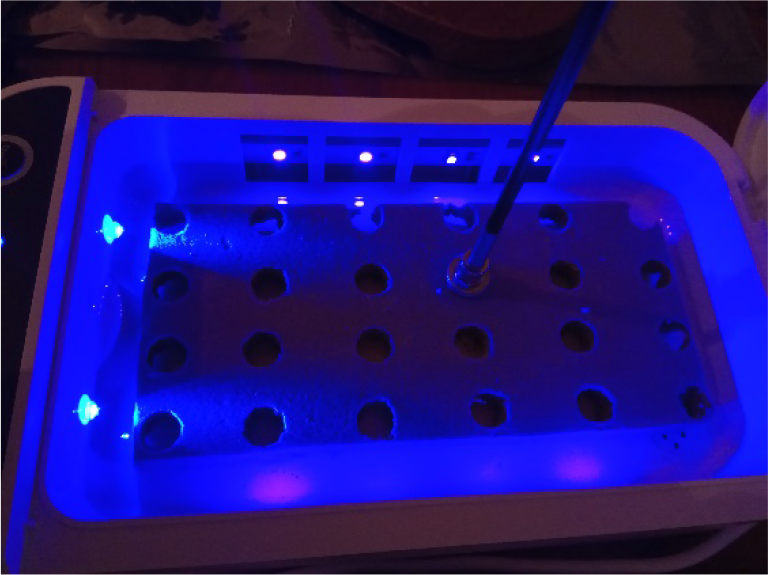
Sanitizing chamber with mobile phone model and spectrum probe.

The measurements were performed with the radiometrically calibrated spectroradiometer, SILVERNOVA from StellarNet. This spectrometer presents a measurement range between 193nm and 1100nm. The integration time was adjusted using the SCOPE mode, to avoid detector saturation and achieve a range of 45.000 bins at the maximum irradiance.

We performed two different sets of measurements. First group of measurements consisted of measuring the irradiance at the lateral of the mobile phone model. In doing that we defined 12 positions shown in figure 3. The separation between the lateral of the chamber and the probe was 5 mm.

**Fig.3.**
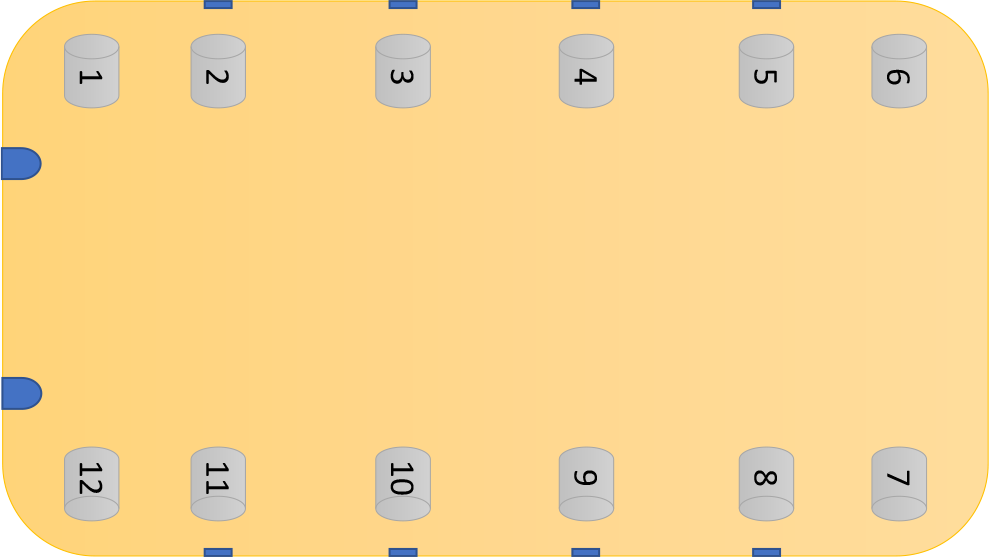
Outline of the lateral irradiance measurements.

We also measured the irradiance at the bottom surface of the mobile phone model. We measured in front of the type 2 LED indicated in fig 1, in the positions shown in figure 4. Measurements in other positions were initially planned but after an initial round of measurements we found that they were not necessary.

**Fig.4.**
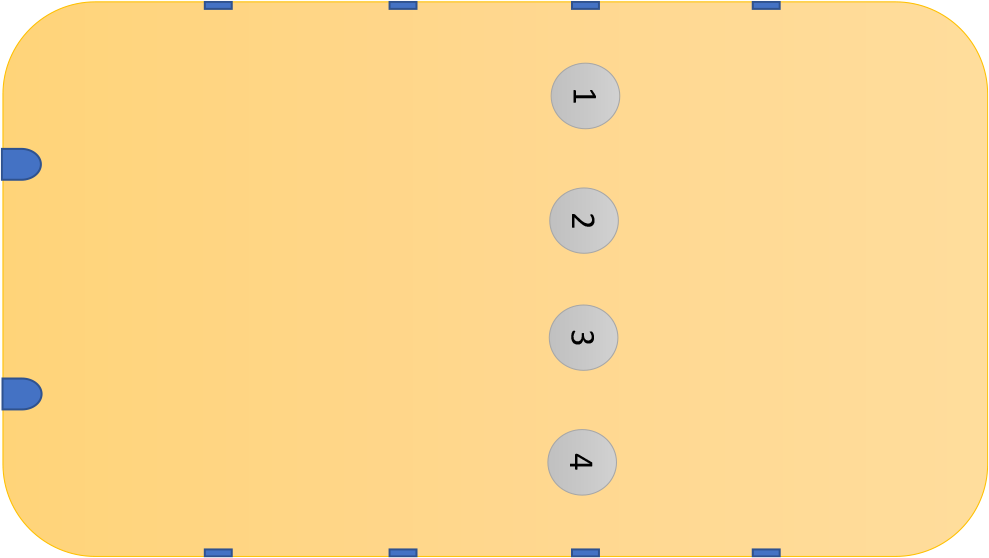
Outline of the irradiance measurements at the bottom of the mobile phone model.

## 3. Results

First, we measured the spectrum of the LEDs, finding that the sanitizing chamber presents three types of LEDs in terms of their emission spectra. In figure 5 we present their measured spectra. Type 1 presents a monomodal emission spectrum with a peak at 397.85 nm and a FWHM of 13.27 nm; Type 2 presents a bimodal emission spectrum with the first maximum at 282 nm with a FWHM of 12.09 nm, and a second maximum at 401 nm with a FWHM of 13.27 nm; Type 3 is a monomodal spectrum with maximum at 464 nm and FWHM of 19.70 nm. So, in the studied disinfection chamber only just one LED emits in the UVC region, and far from the 253 nm emission wavelength clamed in the manual of the device.

**Fig.5.**
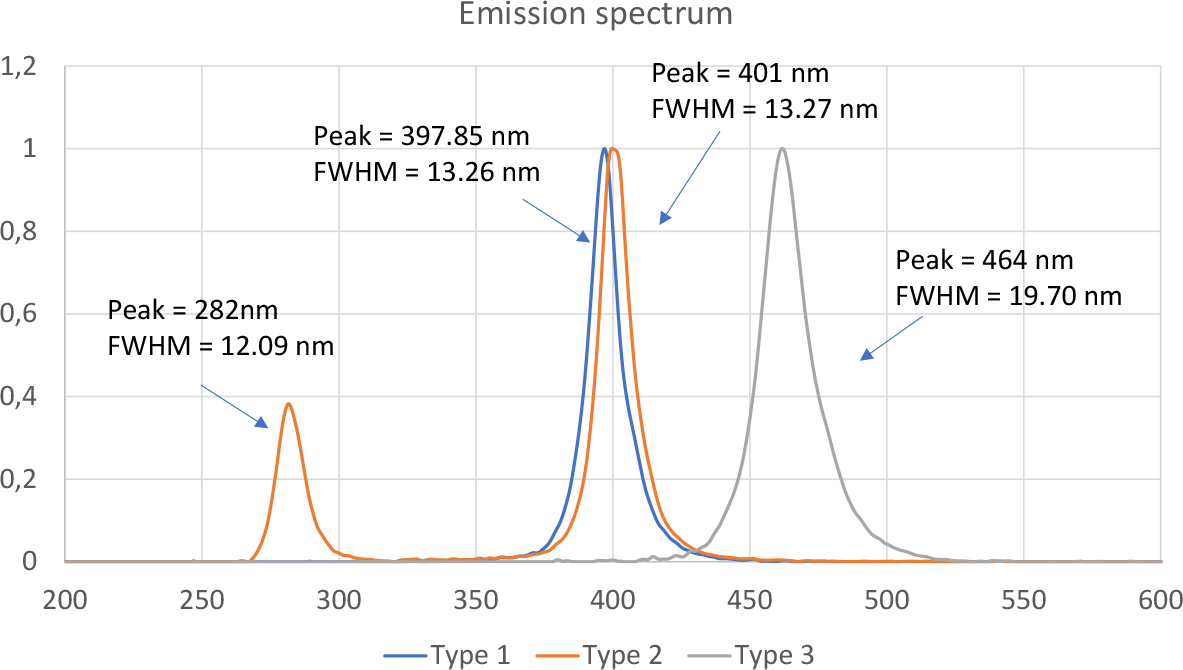
Emission spectra of the three types of LEDs in the disinfection chamber.

In table 1 we present the irradiance measured at the lateral of the mobile phone model at 5mm of the walls of the chamber and in positions indicated in figure 3. We measured the irradiance in three different spectral ranges: first range (250-300) nm containing the first emission lobe of type 2 LED, which have potential biocidal properties; second range blue-violet (380-480)nm; and a third range (250-600) nm containing all the emission spectrum of the three types of LEDs.

**Table 1:** Irradianceμ(W/cm^2^)at 5mm, at the specified wavelength range (nm)

| Position | 1 | 2 | 3 | 4 | 5 | 6 | 7 | 8 | 9 | 10 | 11 | 12 |
| --- | --- | --- | --- | --- | --- | --- | --- | --- | --- | --- | --- | --- |
| 250-300nm | 0 | 10.87 | 19.99 | 1217.61 | 19.3 | 0.11 | 0.04 | 28.44 | 19.37 | 17.32 | 10.88 | 0 |
| **380-480nm** | 101.47 | 1819.31 | 2199.30 | 3396.40 | 2844.09 | 37.66 | 65.17 | 3813.41 | 3280.50 | 3183.55 | 2635.75 | 97.33 |
| **250-600nm** | 101.47 | 1830.18 | 2218.69 | 4614.01 | 2863.39 | 37.77 | 65.21 | 3841.85 | 3299.87 | 3200.87 | 2646.63 | 97.33 |

Figure 6 shows the data of table 1 in a graph of bars in order to better visualize the data.

**Fig.6.**
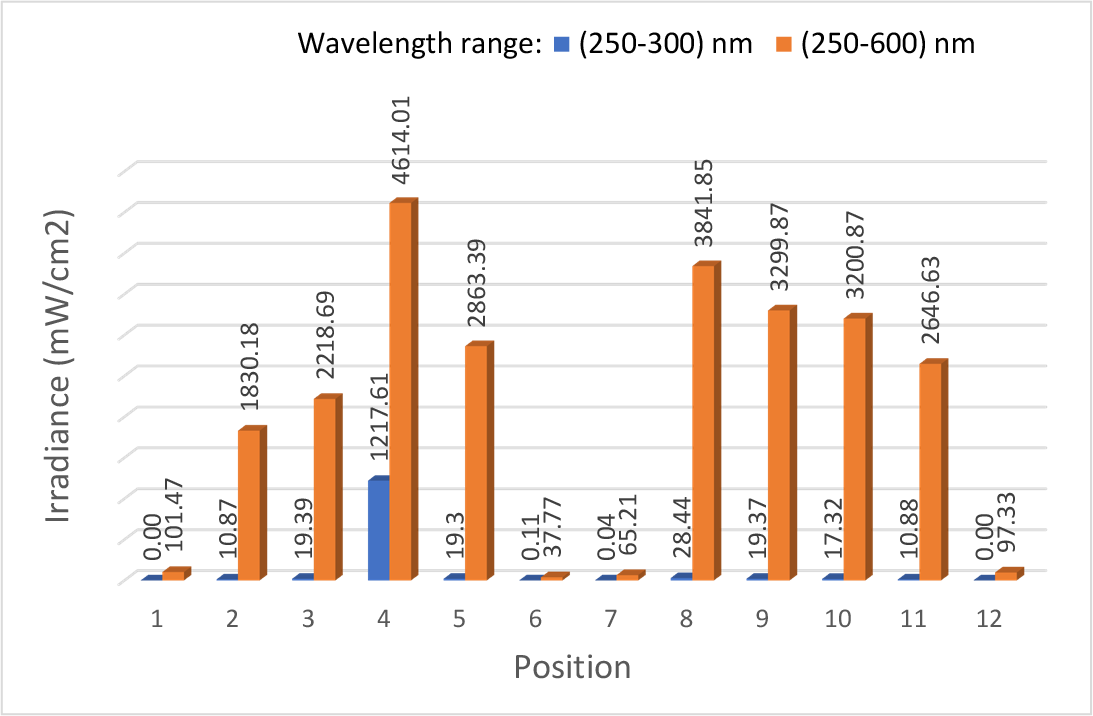
Irradiance measured at the positions described in figure 3, in two wavelength ranges: blue (250,300) nm; red (250, 600) nm.

Table 2 presents the dose achieved in the 5 minutes in which the device is working for disinfection, in the twelve positions indicated in figure 3 and the spectral ranges described before.

**Table 2:** Irradianceμ(W/cm^2^)at 5mm, at the specified wavelength range (nm)

| Position | 1 | 2 | 3 | 4 | 5 | 6 | 7 | 8 | 9 | 10 | 11 | 12 |
| --- | --- | --- | --- | --- | --- | --- | --- | --- | --- | --- | --- | --- |
| **250-300nm** | 0 | 3.26 | 5.82 | 365.28 | 5.79 | 0.03 | 0.01 | 8.53 | 5.81 | 5.19 | 3.26 | 0 |
| **380-480nm** | 30.44 | 545.79 | 659.79 | 1018.92 | 853.227 | 11.29 | 19.55 | 1144.02 | 984.15 | 955.06 | 790.72 | 29.20 |
| **250-600nm** | 30.44 | 549.05 | 665.61 | 1384.20 | 859.02 | 11.33 | 19.56 | 1152.55 | 989.96 | 960.26 | 793.99 | 29.20 |

Table 3 shows the irradiance measured and dose obtained in 5 minutes of exposure at the bottom of the mobile phone model at the positions on figure 4, for the spectral ranges described previously.

**Table 3:**
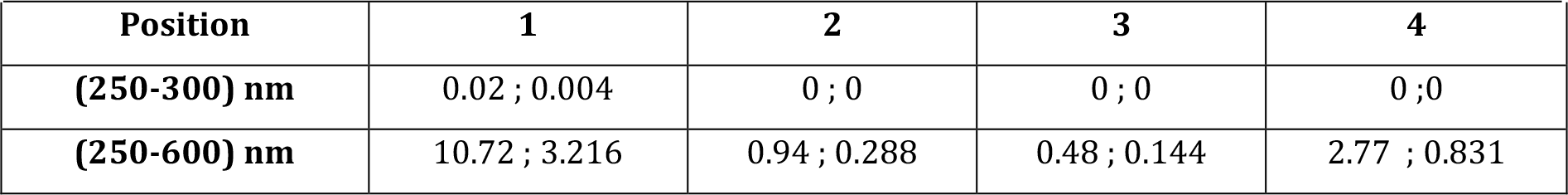
IrradianceμW/cm^2^)at 5mm, at the specified wavelength range (nm)

## 4. Discussion

Covid-19 pandemic has created a new trend in community consumption. Manufacturers and sellers took advantage of the awareness of society with sterilization of objects. Newspapers published and continue to publish frauds concerning sanitizing liquids and devices (as for example face masks which do not protect against the virus). In this work we analyzed a sterilization device which claims to use UVC light at 253.7 nm to sterilize current objects as keys, watches, or mobile phones.

First, we found that the emission wavelength of the LEDs present in the device are far from the 253.7 nm claimed. This wavelength is associated with the emission wavelength of the mercury lamps which are mostly used for sanitization. So, we think that claiming that the device uses this wavelength is just a commercial hook. Just only one of the 8 LEDs supposed to emit UVC light, emits in this range (LED type 2 whose emission spectrum is shown in figure 5). The other 9 LEDs (type 1 and 2) emits in the band region around 400 and 470 nm known as blue-violet region.

Second, we found a high difference in the irradiance and dose achieved at different parts of the sterilizing cabin (see figure 6 and tables 1,2 and 3). Comparing the data of table 1 and 3 we observe that in the irradiance at the cabin in the UVC range is negligible excepting the vicinity of the UVC LED. So, virus disinfection might not be correctly achieved with this device. All studies on disinfection recommend a dose of UVC above 6.7 mJ/cm^2^ for Covid-19 disinfection, and higher for more complex virus [5–10]. Considering the doses achieving during the 5 minutes irradiation, the cabin provides enough dose only in front of the UVC LED. In any other position the dose is not enough for achieving the claimed sterilization. On the other hand, if we face the possibility of disinfecting using UVA, we have to compare the achieved dose, with those recommended in the literature. Reference [10] indicates that with a dose close to 3800 mJ/cm^2^ of UVA is possible to achieve a reduction of 90% of Sars-CoV-2. Reference [11] analyzes other microorganisms, showing for example that a dose of 3600 mJ/cm^2^ in the UVA range is needed for inactivating Escherichia Coli. Besides the work of R. Tomb et.al. [12] on UVA in the range of 380-480 nm shows that inactivation of microorganisms in this range needs more than 4000 mJ/cm^2^. In this range the cabin under study provides a maximum of 1144.02 mJ/cm^2^ in front of LED position 8 at 5mm of the lateral LEDs. So, not enough dose is achieved neither at the UVC range nor at the UVA for sterilizing any object placed inside the cabin.

## 5 Conclusions

The more important conclusion of this work is that we should be aware of what is sell for disinfection. The design of the disinfection cabin has proven to fail, generating an irradiance distribution with very high differences in terms of irradiance, and shadows that do not provide uniform disinfection of the surfaces of the objects placed inside. Additionally, the dose achieved in the 5 minutes of the disinfection treatment is insufficient for disinfection in accordance with the dose recommended in the literature for microorganisms and Sars-Cov-2 inactivation either at the UVC and UVA ranges.

## Data Availability

All data are available in the manuscript

## Acknowledgements

This work was supported by the Spanish Ministry of Economía y Competitividad FIS2016–77319-C2–1-R, Consellería de Educación Program for Development of a Strategic Grouping in Materials – AeMAT Grant No. ED431E2018/ 08 and Xunta de Galicia ref. ED431B2017/64.

